# The influence of repeated mild lockdown on mental and physical health during the COVID-19 pandemic: a large-scale longitudinal study in Japan

**DOI:** 10.1101/2021.08.10.21261878

**Authors:** Tetsuya Yamamoto, Chigusa Uchiumi, Naho Suzuki, Nagisa Sugaya, Eric Murillo-Rodriguez, Sérgio Machado, Claudio Imperatori, Henning Budde

**Author notes:** Corresponding author., *E-mail address:* (T. Yamamoto).

## Abstract

The mental and physical effects of repeated lockdowns are unknown. We conducted a longitudinal study of the influence of repeated mild lockdowns during two emergency declarations in Japan, in May 2020 and February 2021. The analyses included 7,893 people who participated in all online surveys. During repeated mild lockdowns, mental and physical symptoms decreased overall, while loneliness increased and social networks decreased. Subgroup analyses revealed that depression and suicidal ideation did not decrease only in the younger age group (aged 18-29) and that younger and middle-aged people (aged 18-49), women, people with a history of treatment for mental illness, and people who were socially disadvantaged in terms of income had higher levels of mental and physical symptoms at all survey times. Additionally, comprehensive extraction of the interaction structure between depression, demographic attributes, and psychosocial variables indicated that loneliness and social networks were most closely associated with depression. These results indicate that repeated lockdowns have cumulative negative effects on interpersonal interaction and loneliness and that susceptible populations, such as young people and those with high levels of loneliness, require special consideration during repetitive lockdown situations.

---

The onslaught of the coronavirus disease 2019 (COVID-19) remains severe as of June 2021 ^1^. Many countries have implemented lockdowns to prevent the spread of infection. While these lockdowns have been effective in reducing the spread of infection, many negative psychological effects have been reported, including a high prevalence of psychiatric symptoms, such as depression and anxiety ^2–7^.

Previous studies reporting the negative effects during lockdown have been limited to cross-sectional surveys ^3,4,6^ and follow-up studies under single lockdown situations ^5,7^. There have been no longitudinal data demonstrating the effects of repeated lockdowns. Therefore, the question of whether multiple lockdowns have a negative influence on symptoms, or whether the influence can be reduced as people become accustomed to lockdown circumstances remains ^8^. Considering that lockdowns have a significant impact on people’s lives and economic activities ^9^, it can be assumed that repeated lockdowns have different effects on people’s mental health and interpersonal interaction styles than single lockdowns. Since repeated lockdown policies have already been implemented in many parts of the world and may continue to be implemented in the future due to new outbreaks, it is essential to elucidate the influence of repeated lockdowns on mental health.

Japan is one country that has implemented repeated lockdowns. The Japanese government declared a state of emergency on 7 April 2020 and 8 January 2021 and implemented a “mild lockdown” ^6^, an unenforceable request for self-restraint (Figure 1). During the first mild lockdown, a significant increase in psychological distress and depression was reported, despite the lack of legal enforcement of self-restraint ^6,10,11^, the effect of the second mild lockdown on those participants is unclear.

**Figure 1.**
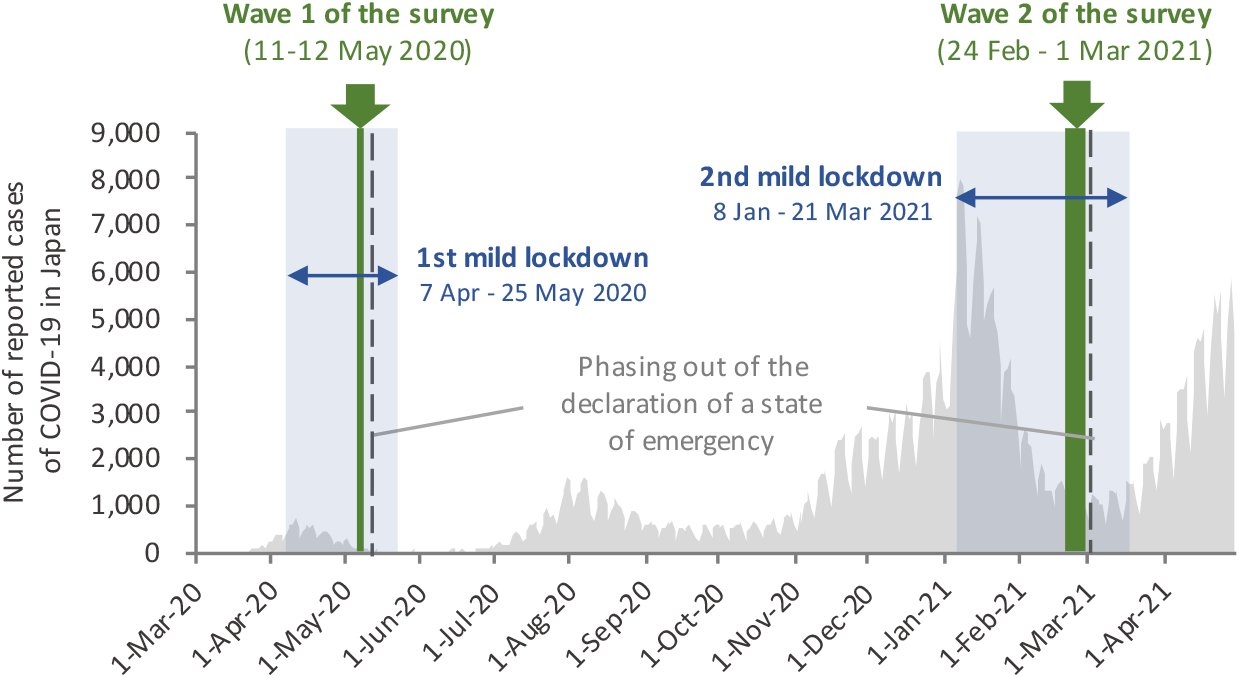
Status of COVID-19 infection in Japan from March 2020 to April 2021 and timing of this study

In this lockdown context in Japan, longitudinal studies of mental health characteristics are critical to understanding the influence of repeated lockdowns. Such analyses will provide important fundamental data for improving lockdown approaches that still permit maximum infection prevention and maintain mental health.

The influence of lockdown is also particularly prevalent among certain groups, such as women, the young, the socially disadvantaged in terms of income, and those with a history of mental illness ^6,12,13^. Because the effects of repeated lockdowns may be specific to these groups, a detailed examination of these associations would provide information regarding what groups require particular attention.

In this study, we examined the influence of repeated lockdowns by conducting a large-scale panel survey of the mental health of people in urban areas under two lockdowns due to declared states of emergency in Japan.

## Results

### Participant characteristics

Of the 11,333 participants in the first wave of the survey, 7,893 participated in the second wave (response rate: 69.6%) (Table 1). The mean age of those who participated in both surveys was 49.6 years (SD=13.7), with the 30-49 age group the most highly represented (41.8%). Of the respondents, 65.6% were married and 68.2% were employed. Compared to those who participated in the first wave only (N = 3,440), those who participated in both were older and had a larger proportion of men and married people (Supplementary Table S1). In addition, those who participated in both experienced greater loneliness and smaller social networks, while fewer people exceeded the cut-off points for depression (PHQ9 ≥ 10) and psychological stress (K6 ≥ 5) and reported suicidal ideation in the first survey (Supplementary Table S1).

**Table 1.** Demographic characteristics of the participants.

| **Characteristics (N=7,893)** | **n (%)** |
| --- | --- |
| **Age, years** |  |
| 18-29 | 637 (8.1%) |
| 30-49 | 3,302 (41.8%) |
| 50-64 | 2,701 (34.2%) |
| ≥ 65 | 1,253 (15.9%) |
| **Gender** |  |
| Men | 4,201 (53.2%) |
| Women | 3,692 (46.8%) |
| **Occupation category** |  |
| Employed | 5,384 (68.2%) |
| Homemaker | 1,236 (15.7%) |
| Student | 111 (1.4%) |
| Unemployed | 901 (11.4%) |
| Other | 261 (3.3%) |
| **Marital status** |  |
| Married | 5,174 (65.6%) |
| Unmarried | 2,719 (34.4%) |
| **Annual household income (JPY)** |  |
| <2 million <sup>a</sup> | 438 (5.5%) |
| 2 million to <4 million | 1,421 (18.0%) |
| 4 million to <6 million | 1,562 (19.8%) |
| 6 million to <8 million | 1,078 (13.7%) |
| ≥8 million | 1,613 (20.4%) |
| Unknown | 1781 (22.6%) |
| **Currently receiving treatment for mental illness** |  |
| Yes | 431 (5.5%) |
| No | 7,462 (94.5%) |
| **Received treatment for mental illness in the past** |  |
| Yes | 875 (11.1%) |
| No | 7,018 (88.9%) |

<sup>a</sup> 2 million JPY = approximately £15,000

### Mental health under repeated mild lockdown in the whole sample

Depression, suicidal ideation, psychological distress, and physical symptoms were significantly lower in the second than the first wave (all ps < 0.001; Table 2). Alternatively, in the second wave, loneliness increased significantly (b = 0.24, 95% CI = 0.15 to 0.33, p < .001) and social network decreased significantly (b = -0.43, 95% CI = -0.54 to -0.33, p < .001). The percentage of patients who exceeded the cut-off score for GAD, measured only in the second wave, was 8.5% (Table 2).

**Table 2.**
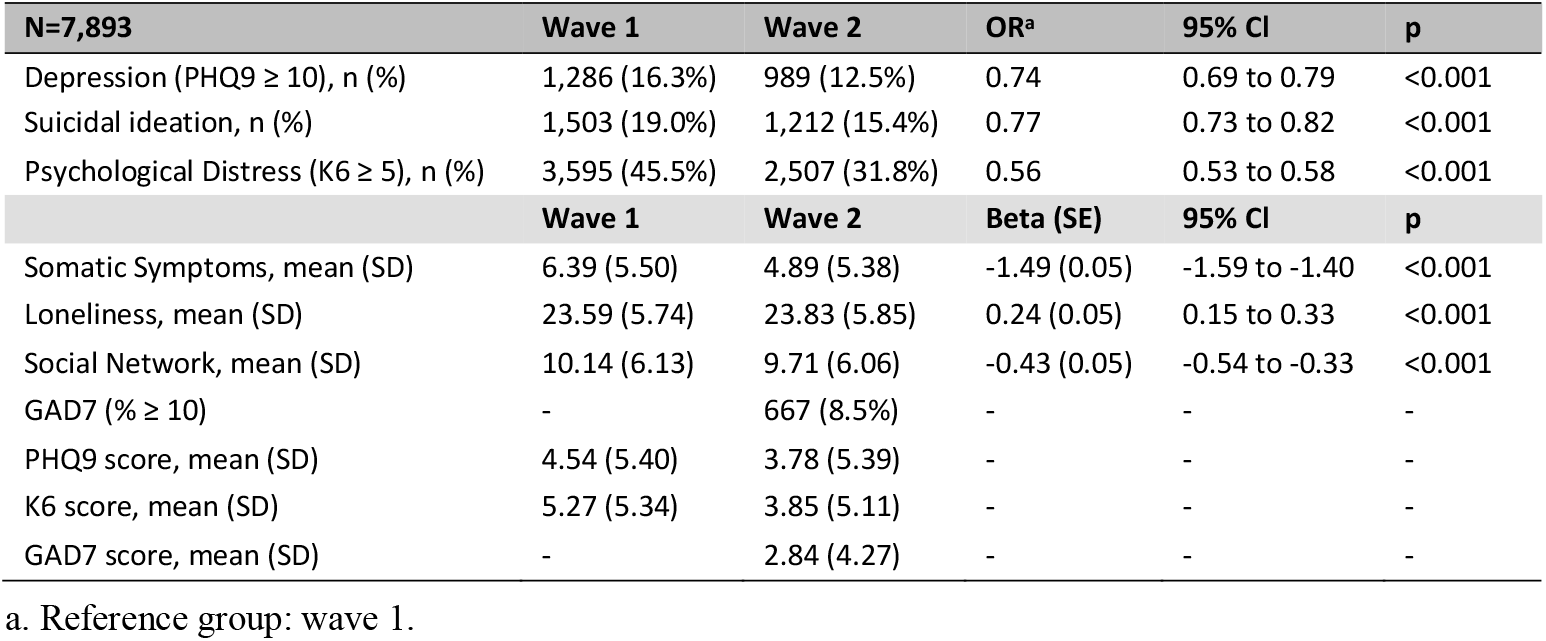
Changes in outcome variables over waves 1 and 2 with odds ratios, betas, and 95% confidence interval.

### Mental health under repeated mild lockdown with a focus on subgroups

The first-and second-wave data for all subgroups are included in the Supplementary Materials (Supplementary Tables S2-S4).

### Estimated rate of depression

There was a significant interaction between gender and wave, and between each age group and wave, for the estimated rate of depression as assessed by the PHQ-9 (Gender: Wald χ2(1) = 7.49, p = .006; Age: χ2(3) = 24.70, p < .001). Both men and women displayed a reduction in depression rate in the second wave (Supplementary Table S2). However, this reduction was significantly smaller among women than men (b = 0.18, 95% CI = 0.52 to 0.32, p < 0.001). Women also had a significantly higher estimated rate of depression in both the first and second waves than men (wave 1: OR = 1.30, 95% CI = 1.15 to 1.46, p < 0.001; wave 2: OR = 1.56, 95% CI = 1.36 to 1.78, p < 0.001). The highest estimated rate of depression from the first to the second wave was observed among 18-29-year-olds, and only this group demonstrated no reduction in the rate of depression (OR = 0.96, 95% CI = 0.73 to 1.27, p = 0.778) (Figure 2, Supplementary Table S2). Additionally, among those aged 65 years and older, the rate of depression was significantly lower in both waves than the other age groups.

**Figure 2.**
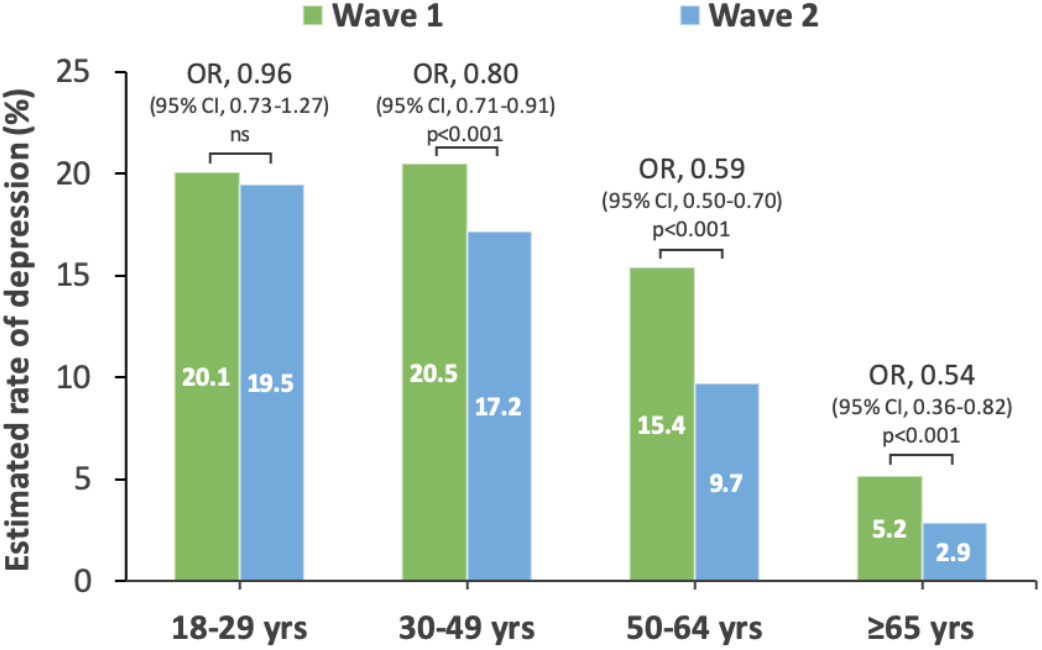
Estimated rate of depression for each age group in wave 1-2. CI, confidence interval; OR, odds ratio

The estimated rate of depression was significantly higher among participants with a history of treatment for mental illness (current, past, current, and past) compared to those without (ORs = 2.58 to 8.53; all ps < 0.001; Supplementary Table S3). The estimated depression rate was significantly higher among participants with an income less than ¥8 million (< ¥2 million, ¥2-4 million, and ¥4-6 million) than those with an income ≥ ¥8 million (ORs = 1.53 to 2.94; all ps < 0.001; supplementary Table S4), and was especially highest among those with an income < ¥2 million.

### Suicidal Ideation

There was a significant interaction between gender and wave, and between each age group and wave, for suicidal ideation (Gender: Wald χ2(1) = 17.87, p < 0.001; Age: χ2(3) = 26.27, p < 0.001). Although suicidal ideation was reduced in the second wave for both men and women (Supplementary Table S2), the degree of reduction in suicidal ideation was significantly smaller among women than men (b = 0.27, 95% CI = 0.14 to 0.39, p < 0.001; Figure 3). Women also reported significantly more suicidal ideation than men in the second wave (OR = 1.29, 95% CI = 1.15 to 1.46, p < 0.001), although this difference was not observed in the first wave (OR = 0.99, 95% CI = 0.89 to 1.11, p = 0.907). Only 18-29-year olds displayed no reduction in suicidal ideation from the first to the second wave (OR = 0.99, 95% CI = 0.76 to 1.29, p = 0.947) (Supplementary Table S2). Additionally, suicidal ideation was significantly lower in both waves among those aged ≥65 than the other age groups (all ps < 0.001).

**Figure 3.**
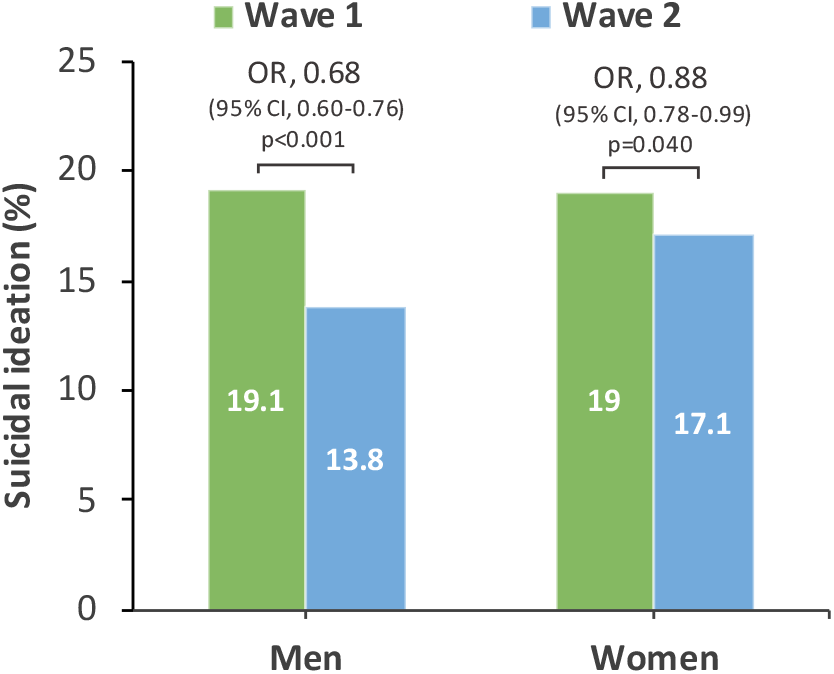
Suicidal ideation by gender in wave 1-2. CI, confidence interval; OR, odds ratio

There were significantly more suicidal ideations among those with a history of treatment compared to those with no history of treatment (ORs=2.38-5.39; all ps < 0.001; Supplementary Table S3), and significantly more among those with an income of <¥8 million compared to those with an income of ≥¥8 million (ORs=1.23 to 2.23; all ps < 0.05; Supplementary Table S4).

### Psychological Distress

There was a significant interaction between age group and wave for the prevalence of stressed ratings according to the K6 (Wald χ2(3) = 19.98, p < .001). All age groups displayed lower stress in the second wave (Supplement S2), but this reduction was significantly smaller among 18-29 and 30-49-year-olds than the 65-year-olds (18-29 years: b = 0.45, 95% CI = 0.21 to 0.69, p < 0.001; 30-49 years: b = 0.32, 95% CI = 0.14 to 0.49, p < 0.001). Among those aged ≥65, stress was significantly lower than in the other age groups in both waves (all ps < 0.001).

Stress was significantly higher among women (OR = 1.55, 95% CI = 1.42-1.70, p < 0.001), those with a history of treatment for mental illness (ORs = 2.79 to 6.87, all ps < 0.001), and those with an income of < ¥8 million (ORs = 1.26 to 1.98, all ps < 0.005) compared to men, those without a history of treatment for mental illness, and those with an income of ≥¥8 million, respectively (Supplementary Tables S2-S4).

### Somatic Symptoms

There was a significant interaction in terms of age and income group (Age: Wald χ2(3) = 33.82, p < 0.001; Income: χ2(4) = 9.70, p = 0.046). Although physical symptoms decreased significantly from the first to the second wave in all age groups (Supplementary Table S2), only 18-29-year-olds demonstrated a significantly smaller decrease in physical symptoms than those aged ≥65 years (b = 0.88, 95% CI = 0.45 to 1.31, p < 0.001). Additionally, in both waves, physical symptoms were reported significantly less frequently among those aged ≥65 years than other age groups (all ps < 0.001). Next, physical symptoms decreased significantly from the first to the second wave for all income groups (Supplementary Table S4), but compared with the group with an income of ≥¥8 million, the group with an income of <¥2 million in the second wave displayed particularly high physical symptoms (b = 1.88, 95% CI = 1.20 to 2.56, p < 0.001), and the groups with an income of 2-4 million and 4-6 million displayed a similarly high trend (¥2-4 million: b = 0.43, 95% CI b 0.07 to 0.80, p = 0.020; ¥4-6 million: b = 0. 44, 95c CI b 0.09 to 0.80, p = 0.015).

Physical symptoms were higher among women (b = 0.87, 95% CI = 0.63 to 1.11, p < 0.001) and those with a history of treatment for mental illness (bs = 3.14 to 6.15, all ps < 0.001) than men and those without a history of treatment for mental illness. (Supplementary Tables S2 and S3).

### Loneliness

A significant interaction was observed for gender and wave (Wald χ2(1) = 13.70, p < 0.001). Among men loneliness did not vary from between waves, (b = 0.09, 95% CI = -0.03 to 0.21, p = 0.162), whereas for women loneliness increased significantly (b = 0.42, 95% CI = 0.07 to 0.55, p < 0.001) (Supplementary Table S2). Loneliness was significantly higher among men than women during both waves (wave1: b = 0.76, 95% CI = 0.51 to 1.01, p < 0.001; wave2: b = 0.43, 95% CI = 0.17 to 0.69, p = 0.001).

Those <65 years of age (bs = 2.40 to 4.55, all ps < 0.001), those with a history of treatment for mental illness (bs = 2.06 to 4.11, all ps < 0.001), and those with an income of < ¥8 million (bs = 1.06 to 4.55, all ps < 0.001) had significantly higher loneliness than those over 65 years of age, those without a history of treatment for mental illness, and those with an income of ≥¥8 million, respectively (Supplementary Tables S2-S4).

### Social Network

A significant interaction was observed for gender and wave regarding social networks (Wald χ2(1) = 4.28, p = 0.039). From the first wave to the second wave, both men and women significantly decreased their social networks (Supplementary Table S2), but the degree of decrease was significantly greater for women than for men (b = -0.43, 95% CI = -0.54- -0.33, p < 0.001). Additionally, in both waves, men had significantly lower social networks than women (wave 1: b = -1.36, 95% CI = -1.62 to -1.09, p < 0.001; wave 2: b = -1.14, 95% CI = -1.40 to -0.87, p < 0.001).

The social network was smaller for those aged 30-64 years (bs = -1.74 to -1.37, all ps < 0.001), those with a history of treatment for mental illness (bs = -1.34 to -2.69, all ps < 0.001), and those with an income of <¥8 million (bs = -1.08 to -5.17, all ps < 0.001) than for those aged ≥65, those without a history of treatment for mental illness, and those with an income of ≥¥8 million (Supplementary Tables S2-S4).

### Exhaustive interaction of factors associated with depression

The final convergence results of the extracted cluster structures are shown in Figure 4 and Supplementary Table S5.

**Figure 4.**
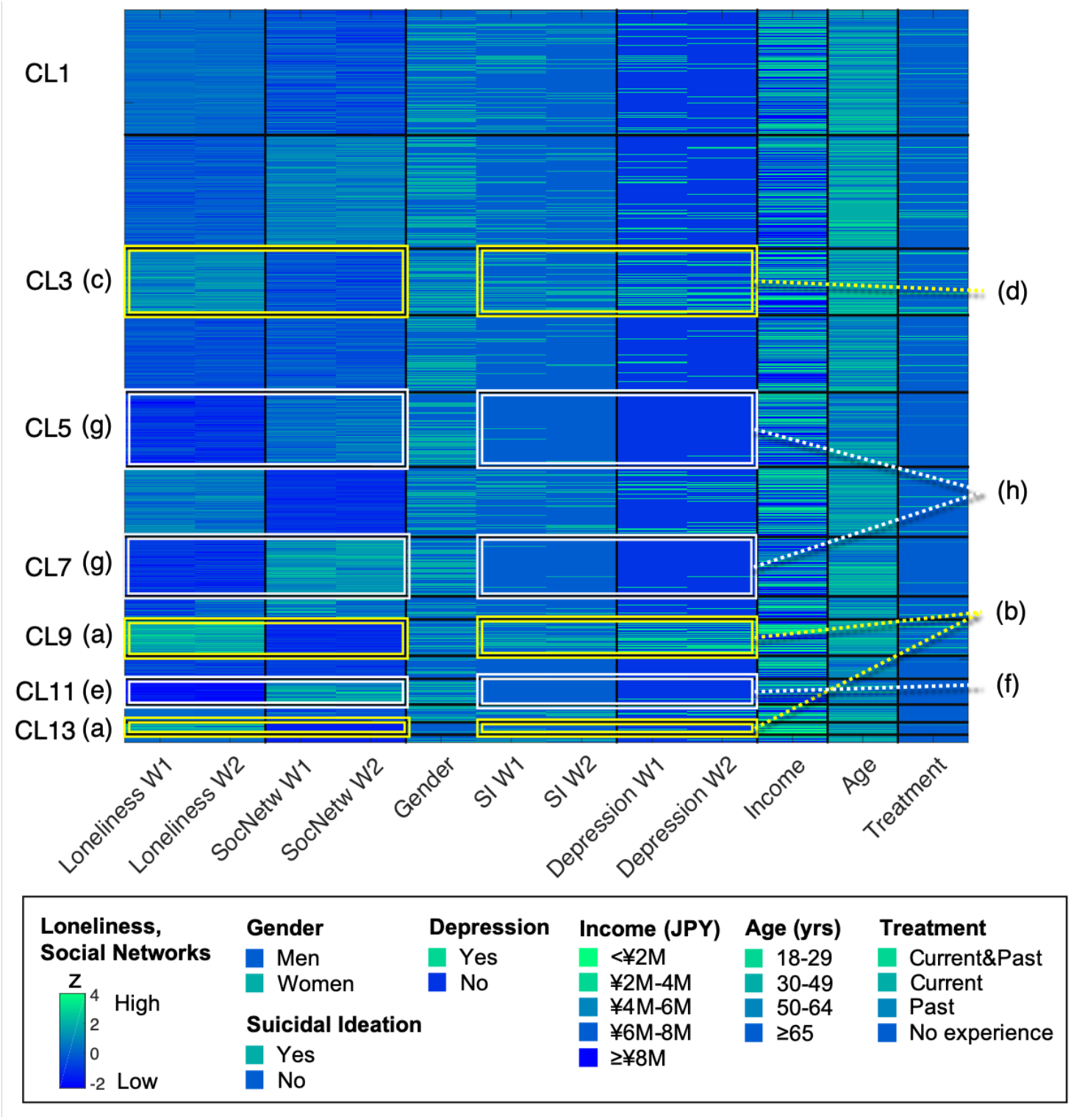
Visualization of an exhaustive interaction structure of factors associated with depression. The rows represent the data of the participants. The columns represent depression, suicidal ideation and their associated factors. The black lines show the clusters of participants and factors respectively. The colours in the cells indicate the magnitude of the z-values or the demographic attributes (see Box at the bottom of the figure). The appended (a), (c), (e), and (g) indicate clusters with significantly higher or lower loneliness and social networks, and (b), (d), (f), and (h) with significantly higher or lower depression and suicidal ideation. M, million; SI, suicidal ideation; SocNetw, social network; Treatment, history of treatment for mental illness; W, wave of the survey

Cluster 9 (CL9, n = 385) and CL13 (n = 136), with significantly higher loneliness and the smallest social network (Figure 4a), had the highest estimated rates of depression in both waves (CL9: 47.8%-54.8%; CL13: 35.3%-42.6%), and both CLs had the highest number of people reporting suicidal ideation (CL9: 48.6%-50.6%; CL13: 40.4%-47.8%)(Figure 4b). Similarly, CL3 (n = 718), the next highest in loneliness (Figure 4c), also showed higher estimated rates of depression (28.6%-37.6%) and suicidal ideation in both waves (30.4%-36.8%)(Figure 4d).

Alternatively, CL11 (n = 281), which had the lowest loneliness and also the most preserved social network (Figure 4e), showed the lowest estimated rates of depression (0.4%-0.7%) and suicidal ideation (both 0.4%) among all CLs in both waves (Figure 4f). Similarly, CL7 (n = 637) and CL5 (n = 794), who were the next least lonely and maintained social networks (Figure 4g), had lower estimated rates of depression (CL7: 1.9%-3.3%; CL5: 2.0%-2.3%) and suicidal ideation (CL7: 2.8%-3.9%; CL5: 2.0%-2.9 %)(Figure 4h).

## Discussion

The results of this study indicate that the overall psychological and physical symptoms decreased during the second mild lockdown compared to the first. During the first mild lockdown due to the declaration of the first ever state of emergency in Japan, there were major economic and social disruption impacts ^14^, which resulted in a significant increase in depression and stress. In contrast, during the second mild lockdown, these symptoms may have been reduced to some extent as a result of adaptation to various environments and life changes caused by the COVID-19 pandemic.

However, at both time points of the study, social network levels indicated social isolation (LSNS-6 < 12) ^15^, which was even more pronounced during the second mild lockdown. Furthermore, loneliness scores increased in the second wave compared to the first wave. These considerations indicate that repeated mild lockdowns cause cumulative changes in interpersonal interactions and that subjective loneliness is maintained and exacerbated.

A striking result when focusing on subgroups was that younger people (18-29 years old) displayed the highest levels of depression and suicidal ideation, and unlike other groups, these indicators did not decrease during the second wave. Additionally, the younger age group demonstrated the smallest decrease in stress and physical symptoms in the second wave of the survey. Women also indicated higher levels of depression, suicidal ideation, and increased loneliness compared to men. These results are consistent with previous reports of deteriorating mental health among young people during the pandemic ^5,7,16^, indicating that younger people are more vulnerable to the negative impacts of repeated mild lockdown. Further, considering that the estimated rate of depression among younger people in this study (22.8%–24%) was significantly greater than that of the general population in Japan in the past (7.9%) ^17^, the present results may indicate that young people experience difficulties with their current situation (e.g. various obstacles related to school and work, financial difficulties due to inability to work), and concerns about the future (e.g. uncertain career paths and employment status) during lockdown situations. Given these considerations, immediate support for young people is highly desirable during repeated lockdown situations. For example, support is needed from educational institutions, companies, and the government by guaranteeing educational and employment opportunities, providing mental health care, and providing information to assuage their concerns.

In addition, those with a history of treatment for mental illness and those who were socially disadvantaged (especially those with incomes <¥2 million) had higher levels of depression, suicidal ideation, stress, physical symptoms, and loneliness, and smaller social networks at both waves. These findings are consistent with previous research ^18,19^ and indicate that these populations are also more susceptible to negative effects of repeated lockdowns. The COVID-19 pandemic has also been reported to cause various economic crises, resulting in lower wages, cutbacks, and general job losses in various industries ^9^, making life increasingly difficult. Therefore, during repeated lockdown situations, a detailed understanding of the difficulties of people with these characteristics is important, and additional social support from social workers and local governments should be considered.

Next, the comprehensive extraction of interactions among variables associated with depression demonstrated that the intensity of loneliness and the size of social networks were most closely related to mental health during both mild lockdown situations. Loneliness and social networks have been considered risk factors associated with mental and physical health ^20,21^, and have been identified as important factors affecting mental health during the COVID-19 pandemic ^4,22^. Considering these previous studies and our results indicating that repeated mild lockdowns cumulatively reduced social networks and maintained or exacerbated loneliness, we believe that the health status of people who display reduced social networks and high loneliness requires special attention in these situations.

### Limitations

In this study, causal factors, such as which approach mitigates or exacerbates the effects of repeated mild lockdowns, have not yet been examined. Therefore, it is necessary to examine the effects of psychosocial variables such as coping strategies and lifestyles in the future.

Additionally, since this study used only an online survey method, the mental health of groups without online access has not yet been examined. Further, since mental health indicators were based on self-reports rather than clinical diagnoses, they may not necessarily correspond to objective assessments by mental health professionals. Therefore, it is necessary to widely use non-online methods in the future to accumulate findings that can be generalised to a larger population.

Finally, those who did not participate in the second wave of the survey had worse mental health compared to those who participated in both waves. Therefore, it should be noted that the present results using only those who participated in both waves may underestimate the impact of repeated lockdowns.

## Conclusion

During the two rounds of mild lockdown in Japan, depression, suicidal ideation, stress, and physical symptoms decreased overall in the second compared to the first wave, while loneliness increased and social networks decreased. These results suggest that repeated mild lockdown exacerbates social isolation and has a cumulative negative effect on loneliness. In addition, depression and suicidal ideation were highest among younger individuals, and as a striking result, did not decrease in the second wave only in the younger age group. Furthermore, women, those with a history of treatment for mental illness, and those from socially disadvantaged backgrounds displayed poorer mental and physical health in both waves. These results indicate that these populations are particularly vulnerable to the negative effects of repeated mild lockdowns. Additionally, loneliness and social networks, in particular, were closely related to mental health at both time points. Given the potential for social isolation and loneliness to be exacerbated by repeated mild lockdowns, these results suggest that during a pandemic situation, people who exhibit these characteristics may require special attention. In the future, it will be essential to conduct continuous research on the health status of potentially vulnerable populations using a combination of various measures, to elucidate protective and risk factors, and to develop support systems tailored to individuals’ difficulties.

## Methods

### Study design, participants, and data collection

This longitudinal study was conducted between 11 May and 12 May 2020 (wave 1) and between 24 February and 1 March 2021 (wave 2). Both surveys were conducted just before the emergency declaration was phased out, and each was set up to assess the psychological impact of a mild lockdown of approximately one month on participants (full elaboration in Figure 1). Participants were recruited by Macromill, Inc. (Tokyo, Japan), a global marketing research company. In the first wave of the survey, approximately 80,000 people were recruited via email. A total of 11,333 people participated in the first wave (the target sample was 11,000 people), and 7,893 participated in the second wave. Participants were recruited only from the seven prefectures where the emergency declaration was first applied (Tokyo, Kanagawa, Osaka, Saitama, Chiba, Hyogo, and Fukuoka prefectures) to sensitively detect the impact of mild lockdown. Since these cities have large populations and a large number of reported cases, it was assumed that they would sensitively reflect the impact of mild lockdown.

The amount of data collected in each prefecture in the first wave was determined according to the ratio of the number of people living in each province to the total population of the seven prefectures (e.g. in the case of Tokyo, 11,428,937 (the population of Tokyo) /46,548,456 (the population of the seven prefectures) ≒24.6%), Tokyo (n = 2,783, 24.6%), Kanagawa (n = 1,863, 16.4%), Osaka (n = 1,794, 15.8%), Saitama (n = 1,484, 13.1%), Chiba Prefecture (n = 1,263, 11.1%), Hyogo (n = 1,119, 9.9%), and Fukuoka (n = 1,027, 9.1%).

The data from the first wave of the survey in this study are part of the dataset used in previous publications ^6,11^, and details of this data can be found in these papers.

A link to the online survey was distributed to those who wished to participate in the study, and the survey was conducted online. All participants completed the anonymous survey voluntarily and provided informed consent online. The online survey form was designed so that all items were required to be answered before proceeding to the next step, thus, there were no missing data in this study. The survey procedures were clearly explained, and participants could interrupt or terminate the survey at any time without providing an explanation. This study was approved by the Research Ethics Committee of the Graduate School of Social and Industrial Science and Technology, Tokushima University (approval number: 212) and has been performed in accordance with the ethical standards laid down in the 1964 Declaration of Helsinki and its later amendments.

### Measures

Depressive symptoms were assessed using the Japanese version of the Patient Health Questionnaire-9 (PHQ-9) ^23^; a score of ≥10 was considered to indicate a high likelihood of depression ^23^ and was used as a cut-off point in this study. Suicidal ideation was assessed using item 9 of the PHQ9 and coded as a binary variable with “not at all” as “no suicidal ideation” and other responses as “suicidal ideation”

Psychological distress was measured using the Japanese version of the K6 ^24^. A threshold of 5 points, commonly used for screening mild to moderate mood/anxiety disorders, was adopted as the cut-off point for psychological distress ^25^. Somatic symptoms were measured using the Japanese version of the Somatic Symptom Scale-8 (SSS-8) ^26^.

Loneliness was measured using the 10-item Japanese version of the UCLA Loneliness Scale Version 3 (UCLA-LS3) ^27^. Social networks and the presence of social support were measured using the Japanese version of the abbreviated Lubben Social Network Scale (LSNS-6) ^15^. LSNS-6 scores <12 indicate social isolation and higher scores indicate a larger social network.

The Japanese version of the Generalised Anxiety Disorder scale-7 (GAD-7) ^28^ was used to assess symptoms of generalised anxiety disorder. Scores above 10 are considered to indicate a moderate level of anxiety ^29^ and were used as a cut-off point in this study. Since the GAD-7 was used only in the second wave, it was used to describe the characteristics of people in the second wave.

These self-report measures are more extensively described in Supplement A in the Supplementary Materials.

Next, to determine the influence of mild lockdowns on groups that have been previously identified as vulnerable to the effects of lockdown ^6,12,13^, demographic attributes such as age, gender, and income were collected, as well as whether or not individuals were currently being treated for mental illness and whether or not they had a history of treatment for a mental illness.

### Statistical analysis

Analyses were performed on a dataset of 7,893 participants from both waves of the survey. First, comparisons were made using t-tests and chi-square tests for the data in the first wave for those who participated in both surveys (N = 7,893) and those who only participated in the first wave survey (N = 3,440).

Next, for each outcome variable, a generalised estimating model (GEE) was constructed to test for changes in the variable between waves for the whole sample. This approach is applicable to binary variables and variables other than normally distributed, and has excellent analytical precision, even for longitudinal data with high within-subject correlation ^30^. In the GEE approach, binomial logit modelling was used for the binary outcome variables (PHQ-9 cut-off score, suicidal ideation, and K6 cut-off score), and linear Gaussian identity modelling was used for the continuous outcome variables (physical symptoms, loneliness, and social network).

Additionally, GEE models were used to examine differences in changes in outcome variables in subgroups focused on gender (men, women), age (18-29, 30-49, 50-64, ≥65), income (< ¥2 million (approximately £15000), ¥2-4 million, ¥4-6 million, ¥6-8 million, ≥¥8 million), and treatment for mental illness (currently in treatment, past treatment, past treatment and still in treatment, no treatment). For analysis of the whole sample, binary outcome variables were analysed using a binomial logit GEE model, and continuous outcome variables were examined using a linear Gaussian identity GEE model. Significant interactions between each subgroup and wave, or main effects for each subgroup, were reported. When analysing the subgroups of income groups, we excluded 1781 participants who answered that they did not know.

To visualise the exhaustive interaction structure of the variables most closely associated with depression, we used nonparametric Bayesian co-clustering ^31^ for gender, age, income, history of treatment for mental illness, depression, suicidal ideation, loneliness, and social networks. Fifteen thousand iterations based on the Bayesian optimisation principle were performed to calculate the log-peripheral likelihood, which indicates the fit of the model. Finally, the model with the highest log-perimeter likelihood was adopted.

The significance level for all tests was set at α = 0.05, two-tailed. The analysis was performed using Matlab R2017a (Mathworks Inc.) and SPSS (version 22.0; SPSS Japan Inc., Tokyo, Japan).

## Data Availability

All data can be obtained from researchers by contacting the corresponding author.

## Acknowledgements

This work was supported by JSPS KAKENHI (grant number 18K13323, 20K10883 and 21H00949), FY2020 Discretionary Funds for Research Director of Tokushima University, FY2020 Special Research Project on Disaster Prevention, and FY2020-2021 Project for Creative Research of the Faculty of Integrated Science, Tokushima University. The funders had no role in study design, data collection and analysis, decision to publish or preparation of the manuscript.

## Authors’ contributions

Conceived and designed the study: TY, CU, NSuz and NSug. Performed the study: TY, CU, NSuz and NSug. Analyzed the data: TY. Wrote the paper, contributed to and have approved the final manuscript: All authors.

## Supplementary Materials

**Supplementary Table S1.** Comparison between those who participated in all surveys and those who participated in wave 1 survey only.

| | Participated in<br>all surveys<br>(n=7,893) | Participated in<br>wave 1 only<br>(n=3,440) | $\chi^2$ or t | df | p | d or V |
| --- | --- | --- | --- | --- | --- | --- |
| Age, mean (SD) | 49.56 (13.71) | 38.66 (13.55) | 39.3 | 6,621 | < .001 | 0.80 |
| Women, n (%) | 3,692 (46.8%) | 2,250 (65.4%) | 333.4 | 1 | < 0.001 | 0.172 |
| Married, n (%) | 5,174 (65.6%) | 1,869 (54.3%) | 128.2 | 1 | < 0.001 | 0.106 |
| Depression (PHQ9 ≥ 10), n (%) | 1,286 (16.3%) | 748 (21.7%) | 48.3 | 1 | < 0.001 | 0.065 |
| Suicidal ideation, n (%) | 1,503 (19.0%) | 754 (21.9%) | 12.4 | 1 | < 0.001 | 0.033 |
| Psychological Distress (K6 ≥ 5), n (%) | 3,595 (45.5%) | 1,854 (53.9%) | 66.9 | 1 | < 0.001 | 0.077 |
| Somatic Symptoms, mean (SD) | 6.39 (5.50) | 7.37 (5.79) | 8.49 | 6,261 | < 0.001 | 0.18 |
| Loneliness, mean (SD) | 23.59 (5.74) | 23.17 (5.59) | 3.60 | 6,717 | < 0.001 | 0.07 |
| Social Network, mean (SD) | 10.14 (6.13) | 11.53 (6.15) | 11.1 | 6,528 | < 0.001 | 0.23 |

PHQ9, Patient Health Questionnaire-9; K6, Kessler Psychological Distress Scale-6.

**Supplementary Table S2.** Changes in outcome variables by age group and gender.

|  | Wave 1 |  |  |  |  | Wave 2 |  |  |  |  |
| --- | --- | --- | --- | --- | --- | --- | --- | --- | --- | --- |
|  | 18-29 yrs | 30-49 yrs | 50-64 yrs | ≥65 yrs | Total | 18-29 yrs | 30-49 yrs | 50-64 yrs | ≥65 yrs | Total |
| **Men (n=4,201)** |  |  |  |  |  |  |  |  |  |  |
| Depression (PHQ9 ≥ 10), n (%) | 25 (18.2%) | 295 (20.7%) | 242 (14.0%) | 53 (5.8%) | 615 (14.6%) | 34 (24.8%) | 224 (15.7%) | 148 (8.6%) | 25 (2.7%) | 431 (10.3%) |
| Suicidal ideation, n (%) | 32 (23.4%) | 366 (25.7%) | 333 (19.3%) | 71 (7.8%) | 802 (19.1%) | 31 (22.6%) | 293 (20.6%) | 219 (12.7%) | 36 (3.9%) | 579 (13.8%) |
| Psychological Distress (K6 ≥ 5), n (%) | 80 (58.4%) | 731 (51.4%) | 671 (38.9%) | 218 (23.9%) | 1,700 (40.5%) | 63 (46.0%) | 525 (36.9%) | 416 (24.1 %) | 112 (12.3%) | 1,116 (26.6%) |
| Somatic Symptoms, mean (SD) | 5.28 (5.18) | 6.46 (5.76) | 6.07 (5.50) | 5.17 (4.50) | 5.98 (5.40) | 4.26 (4.94) | 5.20 (5.69) | 4.50 (5.31) | 3.18 (3.59) | 4.44 (5.17) |
| Loneliness, mean (SD) | 24.04 (5.94) | 25.16 (5.47) | 24.11 (5.39) | 21.70 (5.30) | 23.94 (5.56) | 24.34 (5.37) | 25.29 (5.69) | 24.12 (5.53) | 21.83 (5.40) | 24.03 (5.69) |
| Social Network, mean (SD) | 11.72 (6.58) | 9.04 (6.09) | 9.02 (6.18) | 10.80 (6.34) | 9.50 (6.26) | 11.03 (6.34) | 8.69 (5.94) | 8.67 (6.23) | 10.60 (6.25) | 9.17 (6.20) |
| GAD7 (% ≥ 10), n (%) | - | - | - | - | - | 23 (16.8%) | 158 (11.1%) | 104 (6.0%) | 16 (1.8%) | 301 (7.2%) |
| PHQ9 score, mean (SD) | 4.99 (5.43) | 5.09 (5.69) | 4.05 (5.27) | 2.32 (3.58) | 4.06 (5.21) | 5.48 (6.21) | 4.42 (5.84) | 2.96 (5.15) | 1.49 (3.17) | 3.22 (5.22) |
| K6 score, mean (SD) | 6.62 (5.67) | 5.78 (5.48) | 4.46 (4.96) | 2.80 (3.48) | 4.62 (5.03) | 5.84 (6.35) | 4.41 (5.48) | 2.94 (4.49) | 1.56 (2.84) | 3.23 (4.78) |
| GAD7 score, mean (SD) | - | - | - | - | - | 3.72 (4.87) | 3.42 (4.72) | 2.19 (3.94) | 1.01 (2.39) | 2.40 (4.10) |
| **Women (n=3,692)** |  |  |  |  |  |  |  |  |  |  |
| Depression (PHQ9 ≥ 10), n (%) | 103 (20.6%) | 383 (20.4%) | 173 (17.8%) | 12 (3.5%) | 671 (18.2%) | 90 (18.0%) | 343 (18.3%) | 114 (11.7%) | 11 (3.2%) | 558 (15.1%) |
| Suicidal ideation, n (%) | 112 (22.4%) | 389 (20.7%) | 174 (17.9%) | 26 (7.7%) | 701 (19.0%) | 112 (22.4%) | 370 (19.7%) | 134 (13.8%) | 17 (5.0%) | 633 (17.1%) |
| Psychological Distress (K6 ≥ 5), n (%) | 282 (56.4%) | 1,010 (53.8%) | 482 (49.5%) | 121 (35.7%) | 1,895 (51.3%) | 234 (46.8%) | 774 (41.2 %) | 325 (33.4%) | 58 (17.1%) | 1,391 (37.7%) |
| Somatic Symptoms, mean (SD) | 6.63 (5.40) | 7.09 (5.75) | 7.03 (5.60) | 5.30 (4.48) | 6.85 (5.58) | 5.52 (5.69) | 5.72 (5.71) | 5.45 (5.62) | 3.35 (3.85) | 5.41 (5.58) |
| Loneliness, mean (SD) | 23.44 (5.85) | 23.88 (5.92) | 22.90 (5.76) | 19.72 (5.15) | 23.18 (5.92) | 24.06 (5.90) | 24.29 (6.06) | 23.30 (5.80) | 19.95 (5.15) | 23.60 (6.02) |
| Social Network, mean (SD) | 11.39 (5.90) | 10.58 (5.83) | 10.51 (5.88) | 12.60 (5.99) | 10.86 (5.90) | 10.78 (5.80) | 9.97 (5.80) | 9.97 (5.79) | 12.52 (5.77) | 10.31 (5.83) |
| GAD7 (% ≥ 10), n (%) | - | - | - | - | - | 59 (11.8%) | 222 (11.8%) | 79 (8.1%) | 6 (1.8%) | 366 (9.9%) |
| PHQ9 score, mean (SD) | 5.91 (5.62) | 5.42 (5.67) | 4.88 (5.61) | 2.56 (3.55) | 5.08 (5.55) | 5.51 (5.60) | 4.97 (5.77) | 3.78 (5.24) | 1.63 (2.93) | 4.42 (5.51) |
| K6 score, mean (SD) | 6.64 (5.69) | 6.41 (5.77) | 5.79 (5.44) | 3.68 (3.75) | 6.02 (5.57) | 5.52 (5.72) | 4.97 (5.56) | 4.10 (5.18) | 2.01 (3.15) | 4.54 (5.38) |
| GAD7 score, mean (SD) | - | - | - | - | - | 3.86 (4.57) | 3.82 (4.64) | 2.85 (4.16) | 1.33 (2.38) | 3.34 (4.41) |
| **All (N=7,893)** |  |  |  |  |  |  |  |  |  |  |
| Depression (PHQ9 ≥ 10), n (%) | 128 (20.1%) | 678 (20.5%) | 415 (15.4%) | 65 (5.2%) | 1,286 (16.3%) | 124 (19.5%) | 567 (17.2%) | 262 (9.7%) | 36 (2.9%) | 989 (12.5%) |
| Suicidal ideation, n (%) | 144 (22.6%) | 755 (22.9%) | 507 (18.8%) | 97 (7.7%) | 1,503 (19.0%) | 143 (22.4%) | 663 (20.1%) | 353 (13.1%) | 53 (4.2%) | 1,212 (15.4%) |
| Psychological Distress (K6 ≥ 5), n (%) | 362 (56.8%) | 1,741 (52.7%) | 1,153 (42.7%) | 339 (27.1%) | 3,595 (45.5%) | 297 (46.6%) | 1,299 (39.3%) | 741 (27.4%) | 170 (13.6%) | 2,507 (31.8%) |
| Somatic Symptoms, mean (SD) | 6.34 (5.38) | 6.82 (5.76) | 6.41 (5.55) | 5.20 (4.50) | 6.39 (5.50) | 5.25 (5.56) | 5.50 (5.71) | 4.85 (5.44) | 3.23 (3.66) | 4.89 (5.38) |
| Loneliness, mean (SD) | 23.59 (5.74) | 24.43 (5.77) | 23.68 (5.56) | 21.17 (5.33) | 23.59 (5.74) | 24.12 (5.79) | 24.72 (5.93) | 23.83 (5.64) | 21.32 (5.39) | 23.83 (5.85) |
| Social Network, mean (SD) | 11.46 (6.05) | 9.92 (5.99) | 9.55 (6.11) | 11.29 (6.32) | 10.14 (6.13) | 10.83 (5.87) | 9.42 (5.89) | 9.14 (6.11) | 11.12 (6.18) | 9.71 (6.06) |
| GAD7 (% ≥ 10), n (%) | - | - | - | - | - | 82 (12.9%) | 380 (11.5%) | 183 (6.8%) | 22 (1.8%) | 667 (8.5%) |
| PHQ9 score, mean (SD) | 5.71 (5.58) | 5.28 (5.68) | 4.35 (5.41) | 2.38 (3.57) | 4.54 (5.40) | 5.50 (5.73) | 4.73 (5.81) | 3.26 (5.20) | 1.53 (3.11) | 3.78 (5.39) |
| K6 score, mean (SD) | 6.64 (5.68) | 6.14 (5.65) | 4.94 (5.18) | 3.04 (3.58) | 5.27 (5.34) | 5.59 (5.86) | 4.73 (5.53) | 3.36 (4.78) | 1.68 (2.93) | 3.85 (5.11) |
| GAD7 score, mean (SD) | - | - | - | - | - | 3.83 (4.63) | 3.65 (4.68) | 2.43 (4.04) | 1.09 (2.39) | 2.84 (4.27) |

PHQ9, Patient Health Questionnaire-9; K6, Kessler Psychological Distress Scale-6; GAD7, Generalized Anxiety Disorder Scale-7.

**Supplementary Table S3.** Changes in outcome variables by history of treatment for mental illness.

|  | Wave 1 |  |  |  | Wave 2 |  |  |  |
| --- | --- | --- | --- | --- | --- | --- | --- | --- |
|  | Current<br>(n=118) | Past<br>(n=562) | Current & Past<br>(n=313) | No<br>(n=6,900) | Current<br>(n=118) | Past<br>(n=562) | Current & Past<br>(n=313) | No<br>(n=6,900) |
| **N=7,893** |  |  |  |  |  |  |  |  |
| Depression (PHQ9 ≥ 10), n (%) | 66 (55.9%) | 156 (27.8%) | 170 (54.3%) | 894 (13.0%) | 46 (39.0%) | 128 (22.8%) | 152 (48.6%) | 663 (9.6%) |
| Suicidal ideation, n (%) | 59 (50.0%) | 176 (31.3%) | 159 (50.8%) | 1,109 (16.1%) | 51 (43.2%) | 149 (26.5%) | 149 (47.6%) | 863 (12.5%) |
| Psychological Distress (K6 ≥ 5), n (%) | 98 (83.1%) | 374 (66.5%) | 250 (79.9%) | 2,873 (41.6%) | 83 (70.3%) | 283 (50.4%) | 219 (70.0%) | 1,922 (27.9%) |
| Somatic Symptoms, mean (SD) | 11.47 (5.63) | 8.98 (5.56) | 11.98 (6.91) | 5.83 (5.17) | 9.54 (6.47) | 7.09 (6.00) | 10.83 (7.57) | 4.37 (4.94) |
| Loneliness, mean (SD) | 26.28 (5.26) | 25.29 (5.90) | 27.34 (6.37) | 23.23 (5.61) | 26.69 (5.70) | 25.63 (6.00) | 27.52 (6.57) | 23.46 (5.71) |
| Social Network, mean (SD) | 7.78 (5.48) | 9.04 (5.50) | 7.68 (5.34) | 10.38 (6.18) | 7.61 (5.83) | 8.58 (5.27) | 7.46 (5.53) | 9.94 (6.11) |
| GAD7 (% ≥ 10), n (%) | - | - | - | - | 39 (33.1%) | 89 (15.8%) | 113 (36.1%) | 426 (6.2%) |
| PHQ9 score, mean (SD) | 11.03 (6.56) | 7.11 (5.80) | 11.04 (7.03) | 3.92 (4.90) | 8.69 (7.08) | 6.14 (6.00) | 10.41 (7.66) | 3.20 (4.86) |
| K6 score, mean (SD) | 11.08 (5.96) | 7.84 (5.78) | 10.71 (6.71) | 4.72 (4.94) | 8.99 (6.50) | 5.99 (5.73) | 9.29 (7.14) | 3.34 (4.67) |
| GAD7 score, mean (SD) | - | - | - | - | 6.68 (5.35) | 4.81 (4.93) | 7.91 (6.40) | 2.38 (3.83) |

PHQ9, Patient Health Questionnaire-9; K6, Kessler Psychological Distress Scale-6; GAD7, Generalized Anxiety Disorder Scale-7.

**Supplementary Table S4.**
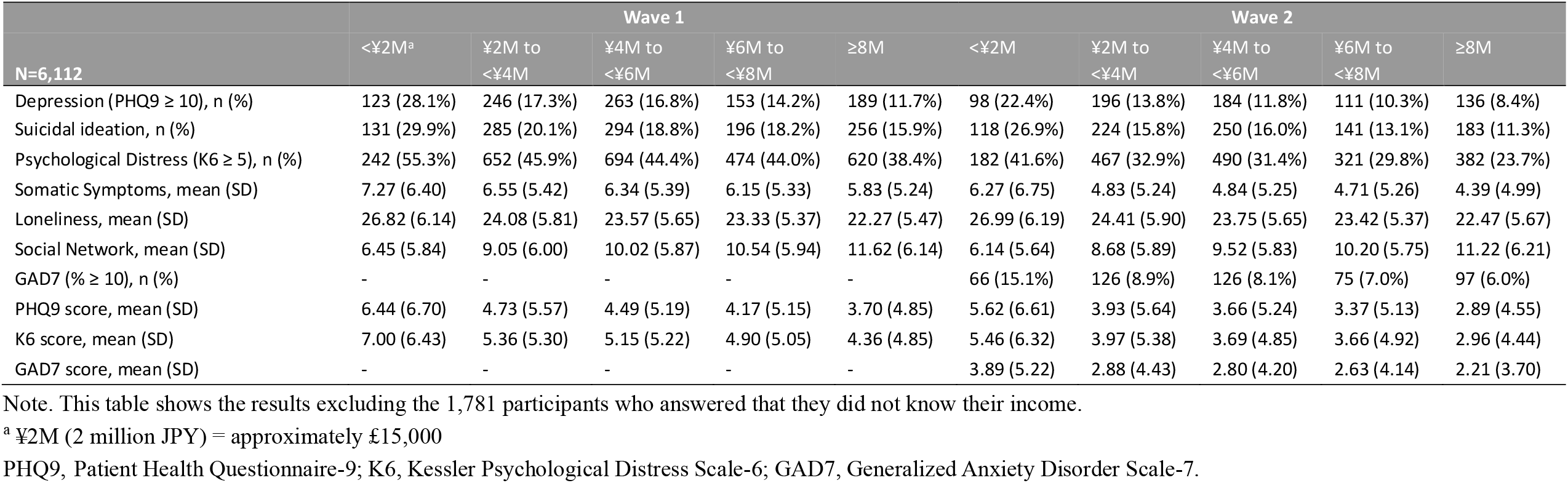
Changes in outcome variables by income group.

|  | Wave 1 |  |  |  |  | Wave 2 |  |  |  |  |
| --- | --- | --- | --- | --- | --- | --- | --- | --- | --- | --- |
|  | <¥2M<sup>a</sup> | ¥2M to<br><¥4M | ¥4M to<br><¥6M | ¥6M to<br><¥8M | ≥8M | <¥2M | ¥2M to<br><¥4M | ¥4M to<br><¥6M | ¥6M to<br><¥8M | ≥8M |
| **N=6,112** |  |  |  |  |  |  |  |  |  |  |
| Depression (PHQ9 ≥ 10), n (%) | 123 (28.1%) | 246 (17.3%) | 263 (16.8%) | 153 (14.2%) | 189 (11.7%) | 98 (22.4%) | 196 (13.8%) | 184 (11.8%) | 111 (10.3%) | 136 (8.4%) |
| Suicidal ideation, n (%) | 131 (29.9%) | 285 (20.1%) | 294 (18.8%) | 196 (18.2%) | 256 (15.9%) | 118 (26.9%) | 224 (15.8%) | 250 (16.0%) | 141 (13.1%) | 183 (11.3%) |
| Psychological Distress (K6 ≥ 5), n (%) | 242 (55.3%) | 652 (45.9%) | 694 (44.4%) | 474 (44.0%) | 620 (38.4%) | 182 (41.6%) | 467 (32.9%) | 490 (31.4%) | 321 (29.8%) | 382 (23.7%) |
| Somatic Symptoms, mean (SD) | 7.27 (6.40) | 6.55 (5.42) | 6.34 (5.39) | 6.15 (5.33) | 5.83 (5.24) | 6.27 (6.75) | 4.83 (5.24) | 4.84 (5.25) | 4.71 (5.26) | 4.39 (4.99) |
| Loneliness, mean (SD) | 26.82 (6.14) | 24.08 (5.81) | 23.57 (5.65) | 23.33 (5.37) | 22.27 (5.47) | 26.99 (6.19) | 24.41 (5.90) | 23.75 (5.65) | 23.42 (5.37) | 22.47 (5.67) |
| Social Network, mean (SD) | 6.45 (5.84) | 9.05 (6.00) | 10.02 (5.87) | 10.54 (5.94) | 11.62 (6.14) | 6.14 (5.64) | 8.68 (5.89) | 9.52 (5.83) | 10.20 (5.75) | 11.22 (6.21) |
| GAD7 (% ≥ 10), n (%) | - | - | - | - | - | 66 (15.1%) | 126 (8.9%) | 126 (8.1%) | 75 (7.0%) | 97 (6.0%) |
| PHQ9 score, mean (SD) | 6.44 (6.70) | 4.73 (5.57) | 4.49 (5.19) | 4.17 (5.15) | 3.70 (4.85) | 5.62 (6.61) | 3.93 (5.64) | 3.66 (5.24) | 3.37 (5.13) | 2.89 (4.55) |
| K6 score, mean (SD) | 7.00 (6.43) | 5.36 (5.30) | 5.15 (5.22) | 4.90 (5.05) | 4.36 (4.85) | 5.46 (6.32) | 3.97 (5.38) | 3.69 (4.85) | 3.66 (4.92) | 2.96 (4.44) |
| GAD7 score, mean (SD) | - | - | - | - | - | 3.89 (5.22) | 2.88 (4.43) | 2.80 (4.20) | 2.63 (4.14) | 2.21 (3.70) |

Note. This table shows the results excluding the 1,781 participants who answered that they did not know their income.
<sup>a</sup> ¥2M (2 million JPY) = approximately £15,000
PHQ9, Patient Health Questionnaire-9; K6, Kessler Psychological Distress Scale-6; GAD7, Generalized Anxiety Disorder Scale-7.

**Supplementary Table S5.**
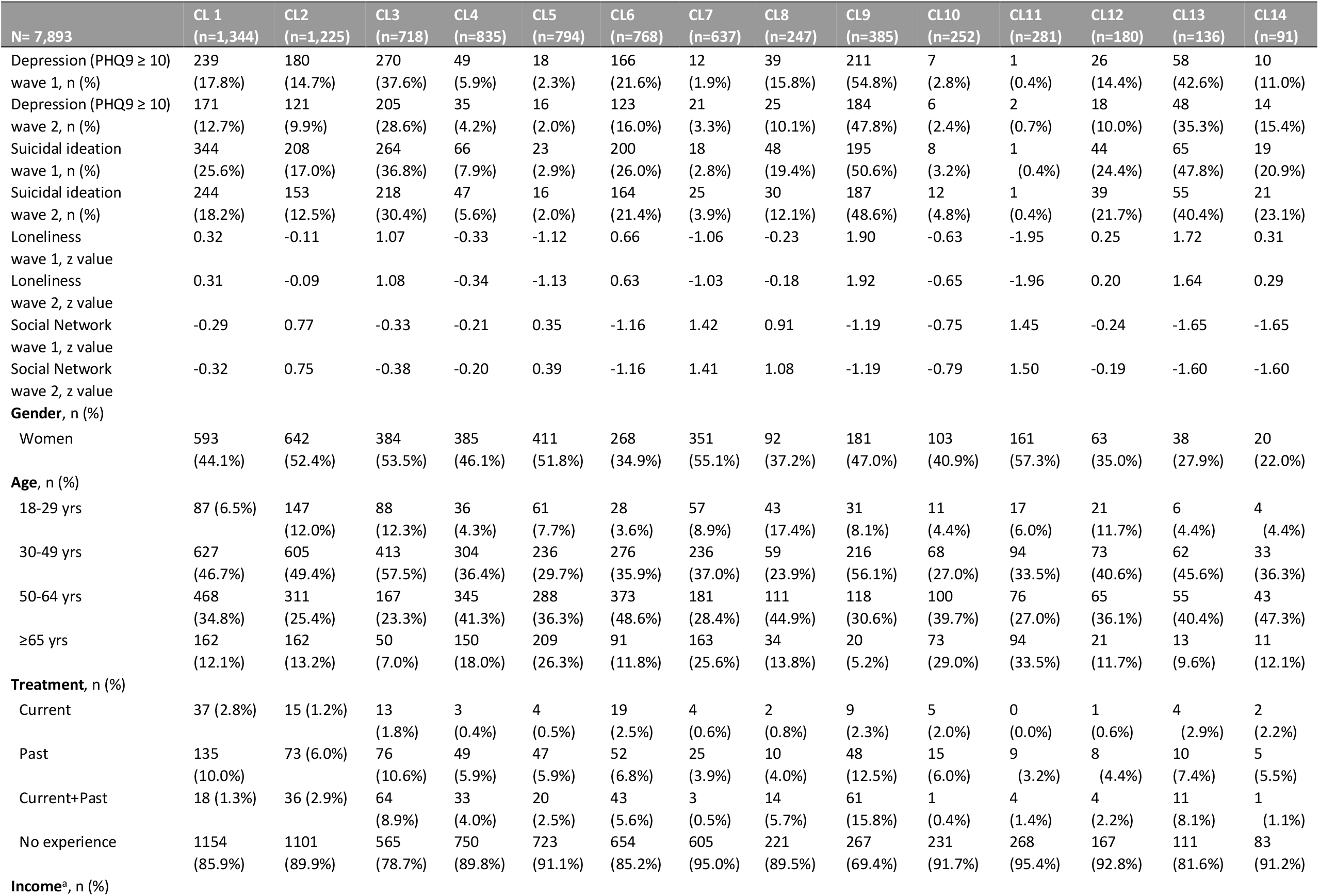
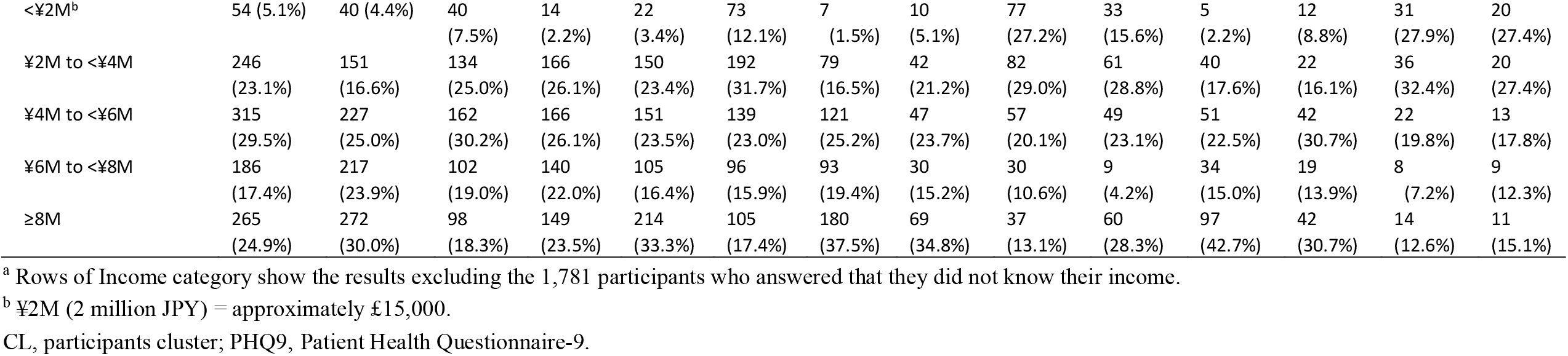
Comprehensive interaction structure of variables associated with depression.

## Supplement A. Full Details of Measures

### Depression and Suicidal Ideation

Depression was measured by the Japanese version of the Patient Health Questionnaire-9 (PHQ-9)(Kumiko Muramatsu et al., 2018). The PHQ-9 consists of nine questions, and participants reported depressive symptoms during the past four weeks assessed by a score of 0 (not at all) to 3 (nearly every day) (Kroenke, Spitzer, & Williams, 2001). We defined a score of ≥10, previously recommended (Kumiko Muramatsu et al., 2018), as a cut-point, meaning that a person is more likely to have major depression. The PHQ-9 is widely used internationally as a screening scale for depression (Siu et al., 2016) with high reliability and validity (Kumiko Muramatsu et al., 2018).

For identifying suicidal ideation (SI), the item 9 of the PHQ-9 was used. The item states, “Over the last 2 weeks, how often have you been bothered by the following problem: thoughts that you would be better off dead, or of hurting yourself in some way?” Answer choices are “not at all”, “several days”, “more than half the days”, or “nearly every day”. SI was coded as a binary variable and considered present for any response other than “not at all”.

### Psychological Distress and Somatic Symptom

Psychological distress was measured by the Japanese version of the K6 (Furukawa, Kessler, Slade, & Andrews, 2003), a six-item screening scale of nonspecific psychological stress in the past 30 days. Each question was rated on a scale of 0 (none of the time) to 4 (all of the time): total scores range from 0 to 24. Given its brevity and high accuracy, the K6 is an ideal scale for screening for mental disorders in population-based health surveys (Furukawa et al., 2003; Kessler, Barker, et al., 2003; Veldhuizen, Cairney, Kurdyak, & Streiner, 2007). We adopted a threshold of five points commonly used to screen for mild-to-moderate mood/anxiety disorders (MMPD) (Prochaska, Sung, Max, Shi, & Ong, 2012). This threshold is the optimal lower threshold cut-point for screening for moderate psychological distress (Prochaska et al., 2012). MMPD was assessed given the risk of progression to more severe disability as well as current distress and disability (Kessler, Merikangas, et al., 2003). Based on 4 years of published data concerning K6 from the Ministry of Health, Labor and Welfare (Ministry of Health Labour and Welfare, 2020), we defined K6 ≥ 5 as ‘psychological distress’.

Somatic symptom burden was assessed by the Japanese version of the Somatic Symptom Scale-8 (SSS-8) (Matsudaira et al., 2016), which consists of 8 items. It consists of questions about common somatic symptoms such as stomach or bowel problems and Headaches in the past week, and requires a response scale of 0 (not at all) to 4 (very much). Scores range from 0 to 32 points. The SSS-8 is highly reliable and valid for assessing somatic symptom burden (Gierk et al., 2014).

### Loneliness and Social Networks

We measured loneliness and social networks using the 10-item Japanese version of the UCLA loneliness scale version 3 (UCLA-LS3) (Arimoto & Tadaka, 2019) and the Japanese version of the abbreviated Lubben Social Network Scale (LSNS-6) (Kurimoto et al., 2011), respectively.

The UCLA-LS3 consists of 10 items, each rated from 1 (never) to 4 (always) (Russell, 1996). The scores range from 10 to 40, with higher scores indicating higher levels of loneliness. The UCLA-LS3 is highly reliable and valid (Arimoto & Tadaka, 2019g) and is internationally used for measuring loneliness (Durak & Senol-Durak, 2010; Shevlin, Murphy, & Murphy, 2015; Zarei, Memari, Moshayedi, & Shayestehfar, 2016). The LSNS-6 consists of three items related to family networks and three items related to friendship networks. The number of people in the network is calculated using a six-point scale (0 = none to 5 = nine or more) for each item (Lubben, 1988). Scores range from 0 to 30 points, with higher scores indicating a larger social network and <12 points indicating social isolation. The LSNS-6 is highly reliable and valid (Kurimoto et al., 2011) and has been used in many countries (Ceria et al., 2001; Martire, Schulz, Mittelmark, & Newsom, 1999; Okwumabua, Baker, Wong, & Pilgram, 1997).

### Anxiety

Anxiety was measured by the Japanese version of the Generalized Anxiety Disorder Scale-7 (GAD-7), a 7-item screening tool. The GAD-7 assesses the frequency with which the seven symptoms of anxiety occurred over the last two weeks (K Muramatsu et al., 2010) by using a scale from 0 (not at all) to 3 (nearly every day): total scores range from 0 to 21. Scores above 10 are considered to indicate a moderate level of anxiety (Spitzer, Kroenke, Williams, & Löwe, 2006) and were used as a cutoff point in this study. The GAD-7 has high reliability and validity for assessing anxiety symptoms (Ruiz et al., 2011).

